# Blood biomarkers in Takotsubo syndrome point to an emerging role for inflammaging in disease pathophysiology

**DOI:** 10.1101/2023.04.05.23288213

**Authors:** M. Nagai, S. Shityakov, M Smetak, H.J. Hunkler, C. Bär, N. Schlegel, T. Thum, C. Y. Förster

## Abstract

Takotsubo syndrome (TTS), an acute cardiac condition characterized by transient wall motion abnormalities mostly of the left ventricle, results in difficulties in diagnosing patients. We set out to present a detailed blood analysis of TTS patients analyzing novel markers to understand the development of TTS. Significant differences in pro-inflammatory cytokine expression patterns, sex steroid and glucocorticoid receptor (GR) expression levels were observed in the TTS patient collective. Remarkably, the measured catecholamine serum concentrations determined from TTS patient blood could be shown to be two orders of magnitude lower than the levels determined from experimentally induced TTS in laboratory animals. Consequently, exposure of endothelial cells and cardiomyocytes in vitro to such catecholamine concentrations did not damage the cellular integrity or function of neither endothelial cells forming the blood brain barrier, endothelial cells derived from myocardium nor cardiomyocytes in vitro. Computational analysis was able to link the identified blood markers, specifically, the pro-inflammatory cytokines and glucocorticoid receptor GR to microRNA (miR) relevant in the ontogeny of TTS (miR-15), and inflammation (miR-21, miR-146a), respectively. Amongst the well-described risk factors of TTS (older age, female gender), inflammaging-related pathways were identified to add to these relevant risk factors or prediagnostic markers of TTS.

## 1. Introduction

Takotsubo syndrome (TTS), also known as broken heart syndrome, stress induced cardiomyopathy, and Takotsubo cardiomyopathy, is an acute cardiac syndrome with rapid onset of chest pain and dyspnea. ^1, 2^ TTS is often triggered by physical and/or emotional stress and is characterized by a transient and reversible severe left ventricular dysfunction which typically recovers spontaneously within hours to weeks.^3^ TTS is a remarkably like acute coronary syndrome (ACS) with almost the same clinical presentations and ST elevations; therefore, differential diagnosis is critically important in the emergency department. The prevalence of TTS has been reported to be approximately 2-3% of all patients with clinical appearance of ACS.^3, 4^

At present TTS-related heart failure is believed to be chiefly induced by a so-called catecholamine (CAT) storm, e.g. increased circulating and myocardial CAT levels with myocardial toxicity.^5–8^ Central to this unifying hypothesis, the abnormal left ventricular contraction pattern is attributed to this CAT surge which triggers left ventricular wall motion abnormality, e.g. widespread dyskinesia in the apical segments and hyperkinesia in the basal segment of left ventricle (LV) with apical ballooning ^3, 4^ ; available information is however controversial ^9–11^: As a pathophysiological mechanism, it is suggested that local differences in left ventricular (LV) adrenergic receptors may underly this LV wall motion abnormarity.^12, 13^ In fact, experimental studies showed that the LV in canine has β2-adrenoceptors (β2-AR) that are expressed much more in the apical than in the basal segments,^14^ the signaling pathways leading to this aetiopathogenesis at any rate remain unclear. Feola et al. further supported this hypothesis with a myocardial PET study showing decreased coronary flow reserve and impaired metabolism in the apical segments during the acute phase of TTS.^15^

Additional possible pathophysiological mechanisms have been suggested such as induced switch in signal trafficking, and autonomic nervous system dysfunction with cardiac sympathetic activation including the mentioned overstimulation of beta receptors.^13, 16–19^ Female gender and age-related lack of female sex hormones are further well known characteristics of TTS patients.^20^

Given that TTS is remarkably like ACS with almost the same clinical presentation and ST elevations; until to date, in the clinical practice, it is difficult to differentiate TTS from ACS at the at the moment of onset. No ECG criteria have been identified that reliably discriminate between TTS and ST- segment elevation myocardial infarction.^21^ The ECG changes are transient and resolve within months in most cases.

Recent studies have identified microRNAs (miRNAs/miRs) as potentially promising sensitive and specific biomarkers for cardiovascular diseases such as TTS, including a unique signature of miR- 1, miR-16, miR-26a and miR-133a, which differentiates TTS patients from STEMI patients.^22, 23^

Using blood samples from real-life patient cohorts, we set out to comprehensively quantify and characterize CAT levels and other blood-borne biomarkers (markers of inflammation, physical/ emotional stress hormone and endogenous sex hormone levels and their receptors) to identify a set of novel TTS-associated biomarkers, pointing to inflammaging-associated triggers for TTS rather than direct CAT effects on cardiomyocytes and endothelial cells of the cardiovascular and cerebrovascular segments. By the help of systems biology we delineate new targets and specific molecular pathways which will allow the development of precision medicine in the prevention and therapy of TTS in the future.

## 2. Materials and Methods

### Study population and human patient serum

Patient serum preparation from human blood was performed as described previously.^24, 25^ Briefly, human blood was collected using S-Monovette tubes (Sarstedt). We sampled blood from 25 pseudonymized patients and subsequently divided into 5 (6 for cytokine measurements) different groups: postmenopausal healthy patients, n=3, premenopausal healthy patients, n=3, ACS acute (<6 weeks), n=4, TTS acute (first 2 weeks after onset), n= 4, TTS subacute (2-6 weeks after onset), n=8, TTS chronic (>6 weeks), n=3. All patients were diagnosed according to the InterTAK Diagnostic Criteria.^26^ As healthy controls subjects were selected that did not present with altered coronary arteries in the coronary angiography diagnostics, while minor cardiovascular or endocrinological diseases were accepted (e.g.: arterial hypertension, diabetes mellitus).

For serum preparation, all the blood was drawn from subjects in using S-Monovette collection tubes (Sarstedt) and incubated at room temperature for 60 min. Then, the blood samples were centrifuged at 1500 × g for 15 min, the serum was isolated, immediately frozen and stored at -80°C until the extraction. All the methods used in this study were performed in accordance with the relevant guidelines and regulations. Serum samples were quantified and analyzed individually.

### Ethics approval and consent to participate

The study protocol was approved by the Hiroshima City Asa Hospital Research Committee (01-3-3) as previously described.^24^

### Study design - Human patient serum

Patient blood serum preparation was performed as described previously.^24, 25^

### Biochemical analyses

Blood samples were drawn after 10-min rest in the supine position in the morning. Adrenaline, noradrenaline, and dopamine concentrations in the plasma were assessed by high-performance liquid chromatography (SRL Inc., Tokyo, Japan). Serum cortisol concentrations were determined by electrochemiluminescence immunoassay (Elecsys®, Roche Diagnostics, Basel, Switzerland). While testosterone was measured using radioimmunoassay kits, 17β-estradiol was measured using an ultra- sensitive radioimmunoassay kit (Elecsys®, Roche Diagnostics, Basel, Switzerland).

### Endothelial cell cultures

The mouse brain capillary endothelial cell line cEND and MyEND were immortalized and isolated as described previously^27–29^ and cultured in Dulbecco’s modified Eagle’s medium (DMEM) with high glucose (Sigma) supplemented with l-glutamine, MEM-vitamin solution, non-essential amino acids (NEA), Na pyruvate, penicillin/streptomycin (P/S) (all from Sigma) 10% heat-inactivated fetal calf serum (FCS) as described previously.^30^

### Human iPSC maintenance and cardiac differentiation

Human induced pluripotent stem cells (iPSC) ^33^ were cultivated in StemMACS medium (StemMACS iPS-Brew XF with supplement, Miltenyi Biotec, #130-104-368) on Geltrex (Gibco, A1413302) and differentiated into cardiomyocytes via Wnt modulation as described before.^34–36^ Briefly, iPSCs were grown to confluency and passaged with Versene (Gibco, 15040066) in StemMACS medium with 2 µM Thiazovivin (Selleckchem, S1459). Directed cardiomyocyte differentiation was initiated at 70- 80% confluency with 5 μM GSK-3 inhibitor XVI (Merck, 361559) in cardio differentiation medium (250 mg human recombinant albumin (Sigma-Aldrich, A9731), 100 mg L-AA (L-ascorbic acid 2- phosphate sesquimagnesium salt hydrate, Sigma-Aldrich, A8960) in 500 ml RPMI medium (RPMI 1640 + GlutaMAX, Gibco, 72400047) for 48 h. Subsequently, medium was changed to cardio differentiation medium supplemented with 5 mM IWP-2 (Selleckchem, S7085), a Wnt signaling inhibitor for 48 h followed by medium changes with cardio differentiation medium every 48 h. At differentiation day 8, when cells start to contract spontaneously, cardiomyocytes were cultured in 1x B-27 (Gibco, 17504001) in RPMI 1640 + GlutaMAX medium, with medium changes every 2 to 3 days.

Cardiomyocytes were purified by metabolic selection^37^ with 4 mM DL-lactate (Merck, L4263, in 1 M HEPES, Carl Roth, HN77.3) in no glucose RPMI medium (RPMI 1640, no glucose, Gibco, 11879020) with 250 mg human recombinant albumin and 100 mg L-AA. Experiments were performed in iPSC-derived cardiomyocytes between day 55-80 of differentiation.

### Catecholamine treatment in vitro

cEND and MyEND were treated with a physiological and supraphysiological CAT mix – corresponding concentration c1-c3 as determined from TTS patient serum, comp. **table 1**, epinephrine (Sigma-Aldrich, E4250), norepinephrine (Sigma-Aldrich, A0937) and dopamine (Sigma-Aldrich, H8502) in the respective cell culture media for 24 h. Epinephrine and norepinephrine were dissolved in 0.5 M HCl, dopamine in medium, for the dilutions medium was used.

**Table 1.**
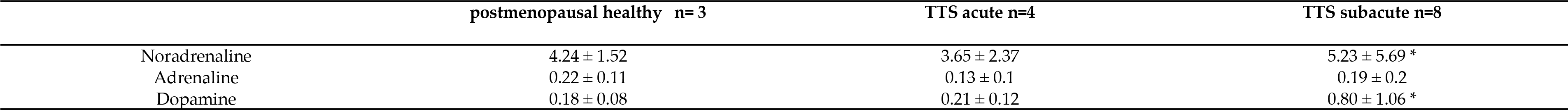
Measured concentration of different CATs (CAT) in serum of postmenopausal healthy patients and patients with TTS in pmol/l ± SD. *: P < 0.05; [TTS acute] = c2, [TTS subacute] = c3 in the following sections Measured concentration of different CATs (CAT) in serum of postmenopausal healthy patients and patients with TTS in pmol/l ± SD. *: P < 0.05; [TTS acute] = c2, [TTS subacute] = c3 in the following sections

For CAT treatment of iPSC-derived cardiomyocytes, human iPSC-derived cardiomyocytes were treated with a physiological mix of these CATs, epinephrine (Sigma-Aldrich, E4250), norepinephrine (Sigma-Aldrich, A0937) and dopamine (Sigma-Aldrich, H8502)), – corresponding concentration c3/ TTS acute levels as determined from TTS patient serum, comp. **table 1** in RPMI medium supplemented with 1x B-27 (Gibco, 17504001) for 24 h. Additionally to the CAT mix designated c3, cells were treated with a supraphysiological concentration of 5 mM isoprenaline (Sigma-Aldrich, I6504).^38^

### Gene expression analysis in cEND and MyEND cells

For RT-PCR, RNA isolation and Real-time RT PCR was performed as described.^30^

### Western blot analysis in cEND and MyEND cells

he images were taken with FluorChem FC2 Multi-imager II (Alpha Innotech). The ImageJ software was used to determine the density of the protein bands.

### Extraction of exosomes from human serum and western blot assay

For extraction of the exosomes, 400 µl of serum was centrifuged (30 minutes, 4°C, 2000 rcf). 300µl of the supernatant was then mixed with 60µl of total exosome isolation reagent (Thermo Fisher Scientific, #4478360) and incubated for 30 minutes at 4°C. After centrifugation (10 minutes, RT, 10000 rcf), the pellet was mixed with 200µl RIPA Buffer containing a protease inhibitor cocktail (ROCHE).

### antibodies

primary: GR H-300 (1:200, Santa Cruz Biotechnology, #sc-8992), mouse anti-claudin-5 (1:1000, Thermo Fisher Scientific, #35-2500), guinea pig anti-occludin (1:100, Acris, #358-504), rabbit anti- ZO-1 (1:1000, Thermo Fisher Scientific, #61-7300), anti-β-actin-HRP (1:25000, Sigma-Aldrich, #A3854), anti-goat VE-cadherin (1:200, Santa Cruz Biotechnology, #sc-6458).

Secondary: goat anti-rabbit HRP (1:15000, System Biosciences, #EXOAB-CD63A-1) was used. and rabbit anti-human CD63 (1:1000, System Biosciences, #EXOAB-CD63A-1).

### Gene expression analysis in cardiomycytes

RNA were isolated with QIAzol Lysis Reagent (Qiagen, 79306) and reverse transcribed with the Biozym cDNA synthesis kit (Biozym, 3314710X) according to manufacturer’s guidelines. Gene expression levels were measured by RT-qPCR with iQ SYBR Green Supermix (Bio-Rad, 1708880). The expression levels of NR4A1 were evaluated with the ΔΔCT method and normalized to the expression of the housekeeper GUSB (β-glucuronidase).

Primer sequences (5’ to 3’):

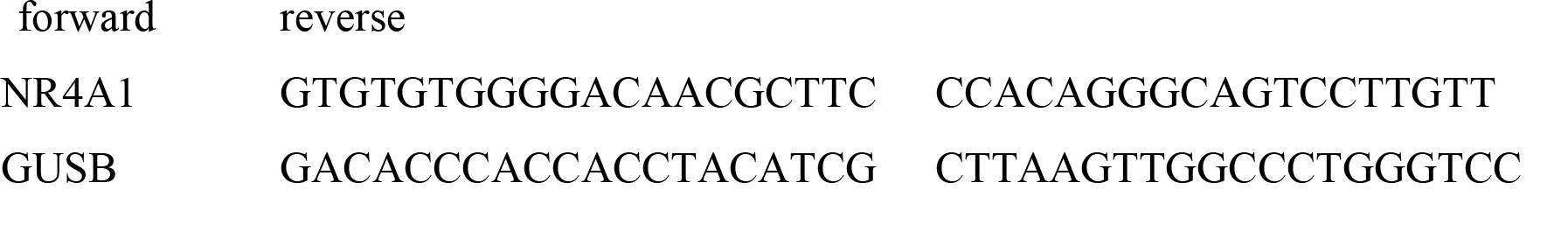

### Cytokine Immunoassay

Cytokines INFγ, IL-2, IL-4, IL-6, IL-10, TNFα, EGF, IL-17, IL-17A, MIP3A, CCL20/MIP3A, RANTES were measured in duplicates in the different patients serum by using a multi-analyte immunoassay (Millipore, HCYTOMAG-60K) with Luminex® bead technology according to manufacturer’s instruction.

### Computational analysis

The Cytoscape v.3.7.1 software was implemented to generate the human gene-miR interaction pathways linked to the searched miRs (miR-15, miR 26a, miR-21, and miR-146a) and proinflammatory cytokines (INFγ, IL-2, IL-4, IL-6, IL-10, TNFα, EGF, IL-17, IL-17A, MIP3A, CCL20/MIP3A, and RANTES). The miRTarBase and TargetScan databases were used for the miR- target gene interaction types together with a list of the searched miRs by using the CyTargetLinker app v4.10.^39^ For the gene-gene interaction network containing proinflammatory cytokines, the GeneMANIA algorithm was applied comprising 911 networks and 24280 genes.^40, 41^ Finally, the merge tool as a part of the Cytoscape pipeline was implemented to merge previously generated networks combining analyzed miRs and cytokines.

### Analysis and statistics

Data are shown as mean and standard deviation (SD) of biological independent experiments. One-way ANOVA with Dunnett’s multiple comparison test or Tukey’s t-Test as indicated in the figure legends was performed to evaluated the statistical significance between untreated und treated cells with GraphPad Prism 8. P < 0.05 was considered as significant, * P < 0.05, ** P < 0.01, *** P < 0.001.

## 3. Results

### Quantification of catecholamines in serum of patients with different heart diseases

The CAT noradrenaline, adrenaline and dopamine were measured in serum of postmenopausal healthy patients and patients admitted with TTS (acute and subacute phase). Significantly elevated noradrenaline (5.23 ± 5.69 pmol/l) vs. (4.24 ± 1.52 pmol/l) and dopamine (0.80 ± 1.06 pmol/l ) vs. (0.18 ± 0.08 pmol/l) levels were detected in the TTS subacute group exclusively, all other levels did not differ significantly from the control group, postmenopausal healthy females (Table 1). Among the three CAT, noradrenaline showed highest absolute serum levels among the groups, while dopamine and adrenaline varied.

In the following, endothelial cells and cardiomyocytes were treated in vitro with the different CAT concentrations as determined from TTS acute and TTS subacute patient serum as opposed to pharmacological concentrations (concentration c1 pharmacological concentrations^42^: 150µM dopamine, 1µM epinephrine, 1µM norepinephrine; c2 postmenopausal healthy control; c3 TTS acute; c4 TTS subacute, comp. Table 1).

### The concentration-dependent effects of CAT on cEND brain endothelial cells in vitro

To investigate the concentration-dependent change in protein expression of barrier-forming proteins claudin-5, occludin, VE-cadherin and ZO-1 following CAT exposure in vitro, cEND cells were incubated with the different concentrations of the CAT dopamine, epinephrine and norepinephrine as determined from patient serum comp. Table 1 compared to an exposure to supraphysiological/ pharmacological concentrations ^42^ for 24h.

Changes in protein levels were determined by Western blot analysis and compared to the untreated control group (Figure 1). Only the supraphysiological/ parmacological c1 concentration showed significant effects on the protein expressions: ZO-1, VE-cadherin and occludin expression were significantly reduced (0.451 ±0.130-fold) and (0.186 ±0.069-fold). Surprisingly, claudin-5 expression was significantly increased (1.761 ±0.312-fold). cEND monolayer exposure to CATs in concentration ranges as as determined from TTS patient serum (c2, c3, c4) showed no significant change in the protein expression.

**Figure 1.**
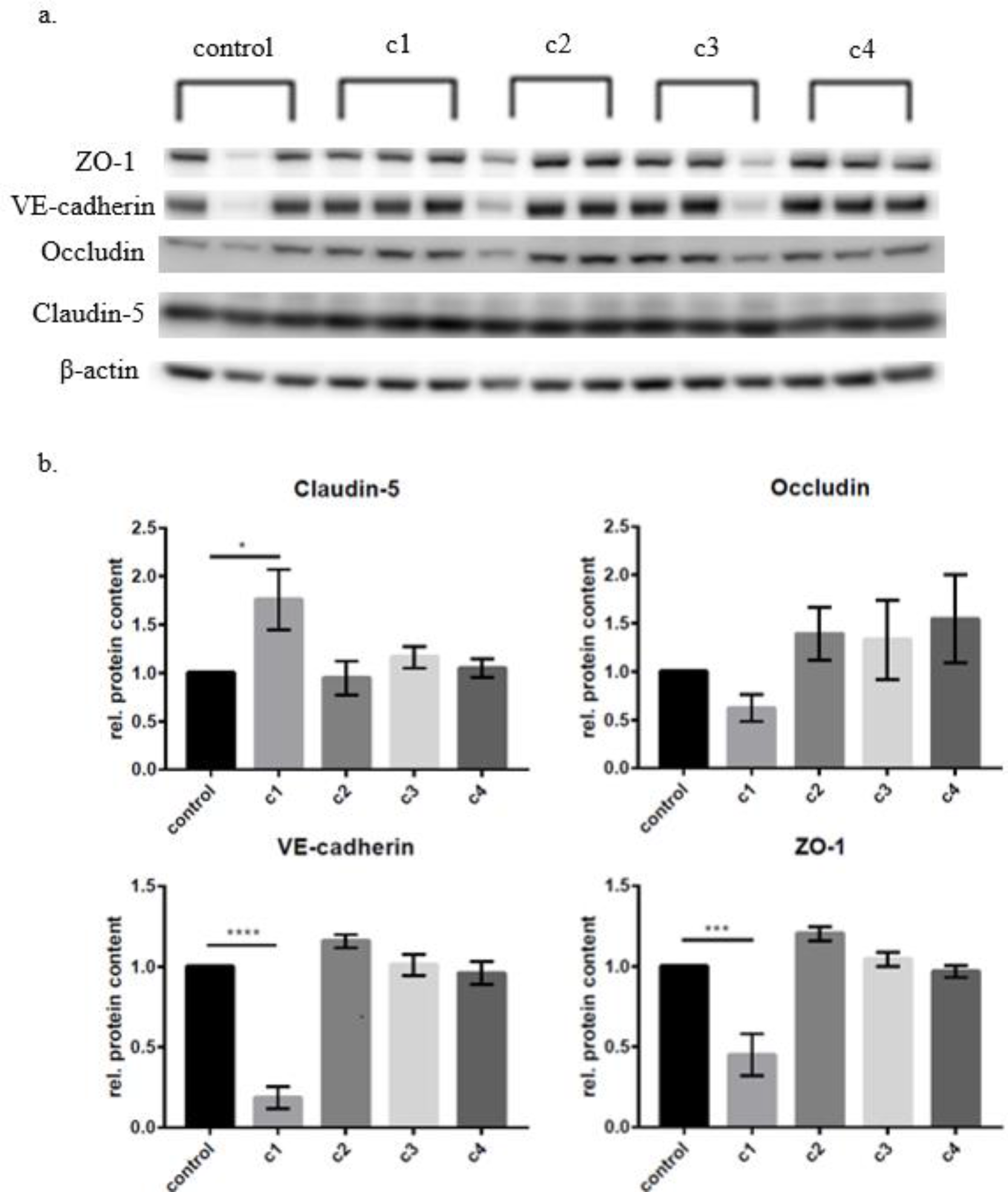
Changes of protein expression in cEND cells after incubation with the CATs norepinephrine, epinephrine and dopamine in different concentration (c1 pharmacological concentrations) ^42^: 150µM dopamine, 1µM epinephrine, 1µM norepinephrine; c2 postmenopausal healthy control; c3 TTS acute; c4 TTS subacute, comp. **table 1**). a) Western blots displaying xpression of the different proteins after 24h of incubation. b) densitometric evaluation. Changes of protein expression was normalized to β-actin. Data are the means (±SEM) of 3 independent experiments. Statistical significance was evaluated using Dunnett’s multiple comparisons test. *: P<0.05; ***: P < 0.001; ****: P < 0.00001

The concentration-dependent changes in mRNA expression were determined by qRT-PCR analysis and compared to the untreated control group (Figure 2). Only the supraphysiological/ parmacological c1 concentration showed significant effects on the mRNA expression. Claudin-5 expression was significantly reduced (0.610 ±0.059-fold). Also, VE-cadherin and ZO-1 showed a significant reduction (0.582 ±0.139-fold) and (0.478 ±0.085-fold). Exposure of cEND monolayers with CATs in concentrations as determined from TTS patient serum (c2, c3, c4) and occludin showed no significant change on the mRNA expression (Figure 2).

**Figure 2.**
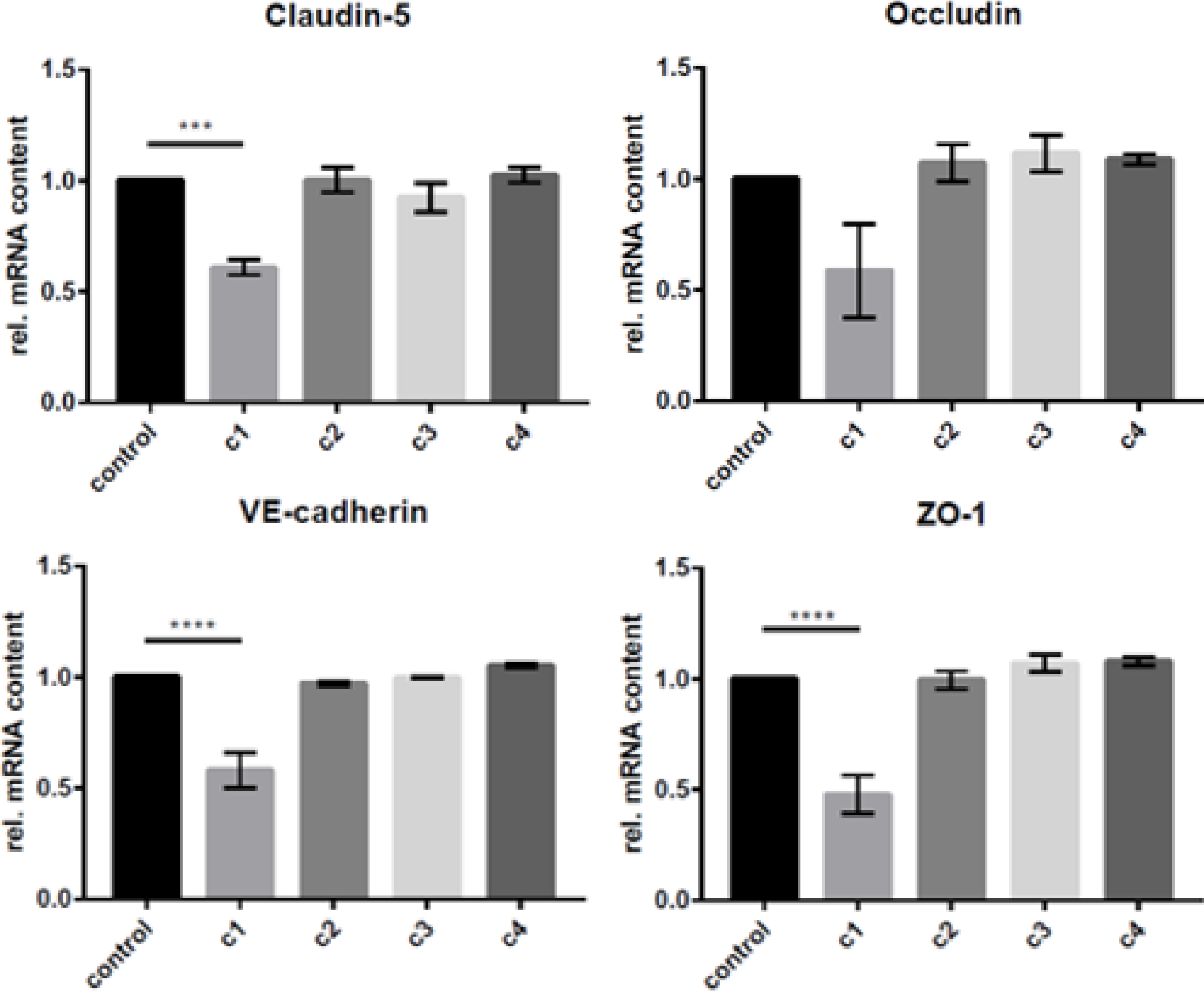
Changes of mRNA expression in cEND cells after incubation with the CATs norepinephrine, epinephrine and dopamine in different concentrations (c1 pharmacological concentrations ^42^: 150µM dopamine, 1µM epinephrine, 1µM norepinephrine; c2 postmenopausal healthy control; c3 TTS acute; c4 TTS subacute, comp. Table 1). Changes of protein expression was normalized to β-actin. Data are the means (±SEM) of 3 independent experiments. Statistical significance was evaluated using Dunnett’s multiple comparisons test. ***: P < 0.001; ****: P < 0.00001

### The concentration-dependent effects of catecholamines on MyEND cells

To investigate the concentration-dependent change in protein expression, MyEND cells were incubated with the different concentrations of the CATs dopamine, epinephrine and norepinephrine as described for Fig. 2 for 24h. Changes in protein levels were determined by Western blot analysis and compared to the untreated control group (Figure 3). Only the supraphysiological/ parmacological c1 concentration showed significant effects on the protein expressions: Claudin-5 expression levels were significantly decreased in this application (0.275 ±0.067-fold), and occludin expression was significantly reduced (0.686 ±0.122-fold). The other CAT concentrations applied as determined from TTS patient serum (c2, c3, c4) showed no significant change in the protein expression of claudin-5 or occluding (data not shown).

**Figure 3.**
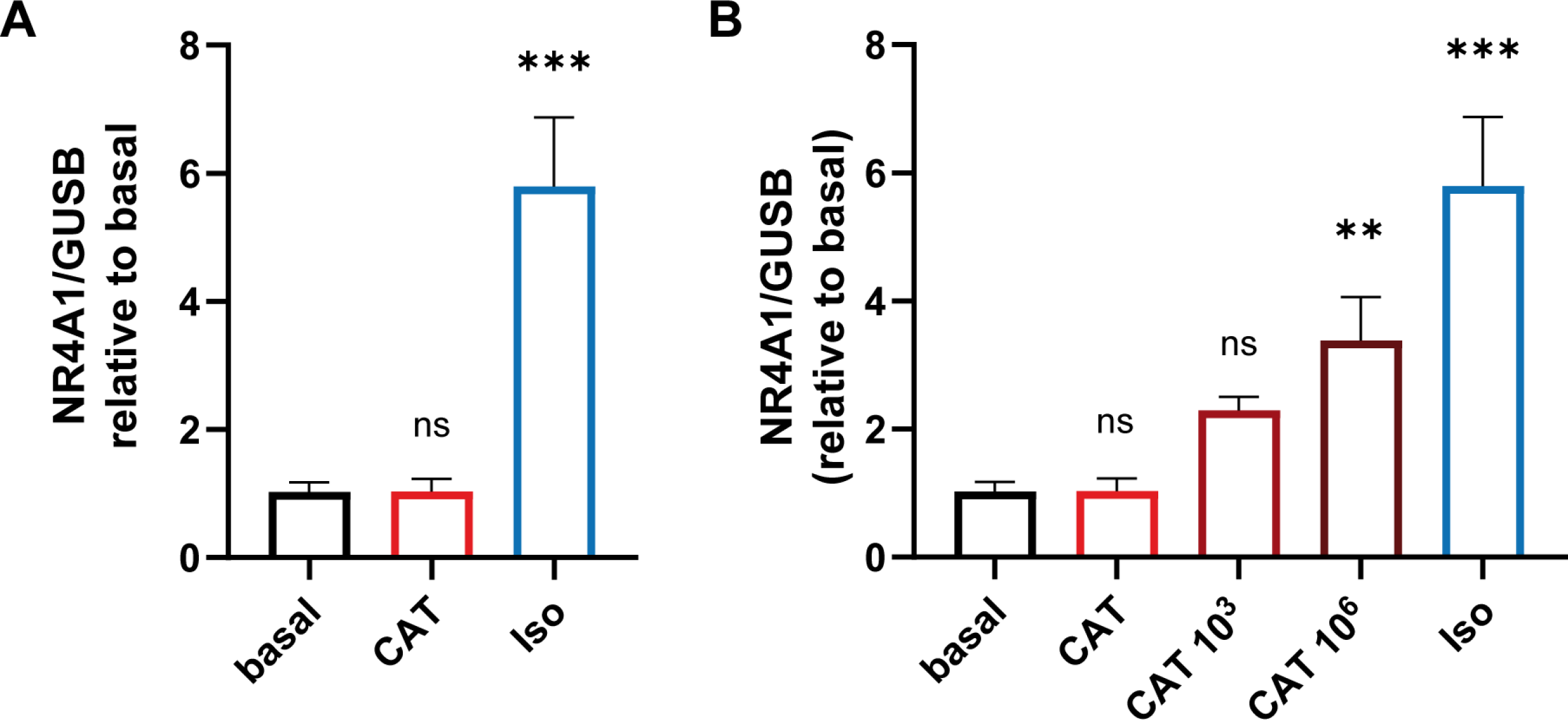
Physiological CAT concentrations did not alter cardiac stress marker NR4A1 in cardiomyocytes. A Human iPSC-derived cardiomyocytes were treated with physiological concentrations of a CAT mix as determined from acute TTS (c3 comp. Table 1) and supraphysiological concentrations of isoprenaline (Iso) for 24 h. The expression level of the cardiac stress marker nuclear receptor subfamily 4 group A member 1 (NR4A1) was analyzed by RT-qPCR, mean ± SD of three independent experiments is shown. B Human iPSC-derived cardiomyocytes were treated with physiological concentrations of CAT as determined from acute TTS (c3) and multiples (10^3^, 10^6^) of it. Expression of NR4A1 was determined by RT-qPCR, mean ± SD of triplicates is shown. Statistical significance was analyzed with one-way ANOVA with Dunnett’s correction in comparison to basal, ** P < 0.01, *** P < 0.001, ns P > 0.05.

The concentration-dependent changes in mRNA expression were equally determined by RT-qPCR analysis and compared to the untreated control group (Figure 4). Claudin-5 (0.312 ±0.09-fold) and occludin (0.473 ±0.055-fold) expression were significantly reduced after incubation with CAT in concentration c1 (pharmacological). Occludin expression was significantly reduced as well after incubation with c1. VE-cadherin and ZO-1 showed a significant reduction after c1 incubation (0.438 ±0.020-fold) and (0.463 ±0.0259-fold), respectively.

**Figure 4.**
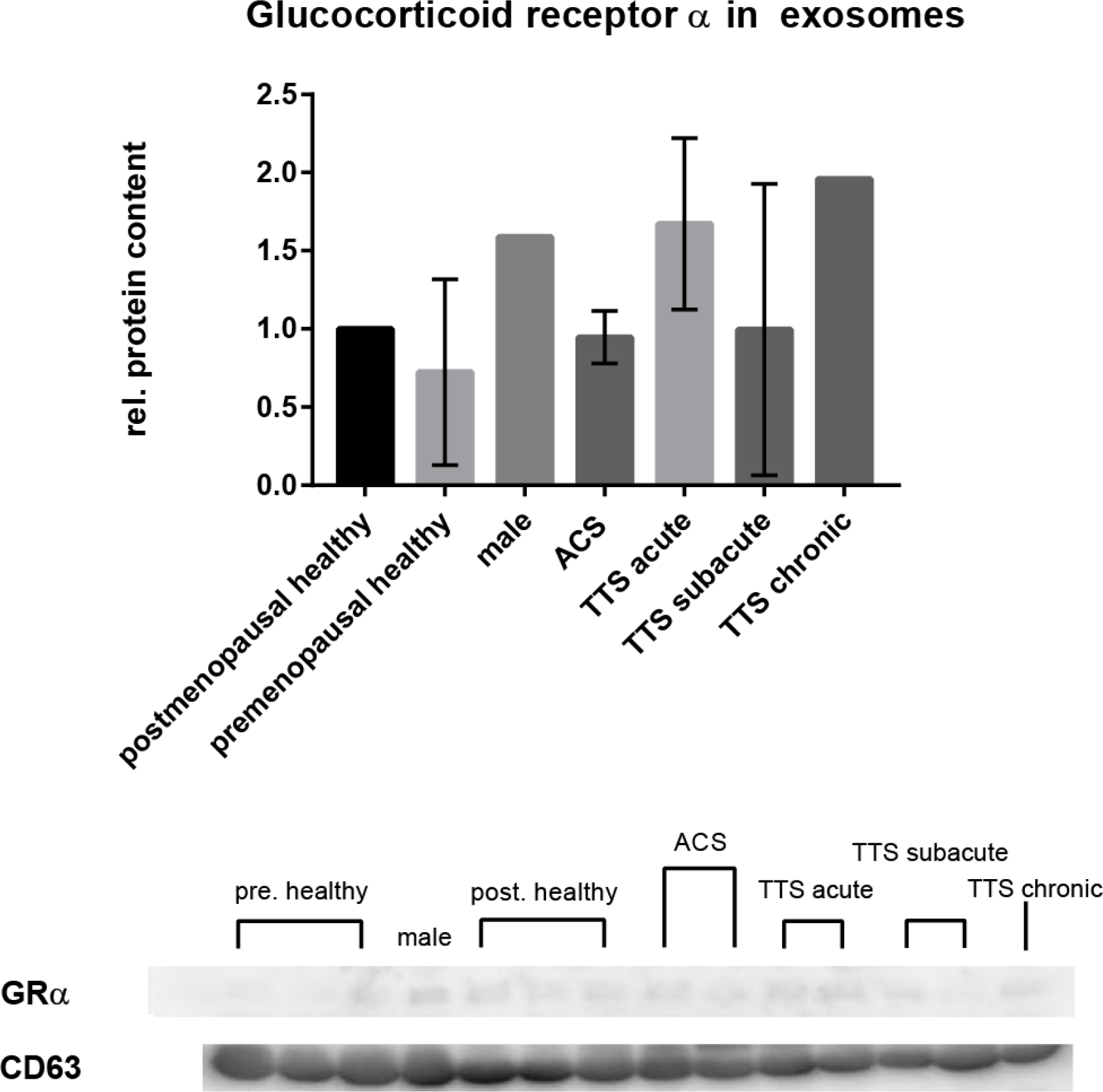
Changes in the protein expression of glucocorticoid receptor α in exosomes from serum from patients with various heart diseases. Expression of glucocorticoid receptor α in the exosomes. (densitometric evaluation) a) Membranes of the Western blots used in this experiment. Glucocorticoid receptor α protein expression was normalized to CD63. Data are the means (±SD) of n independent patients (postmenopausal healthy: n=3, premenopausal healthy: n=3, ACS acute: n=4, TTS acute: n=4, TTS subacute: n=8, TTS chronic: n=3). Statistical significance was evaluated using Tukey’s multiple comparisons test.

**Figure 5.**
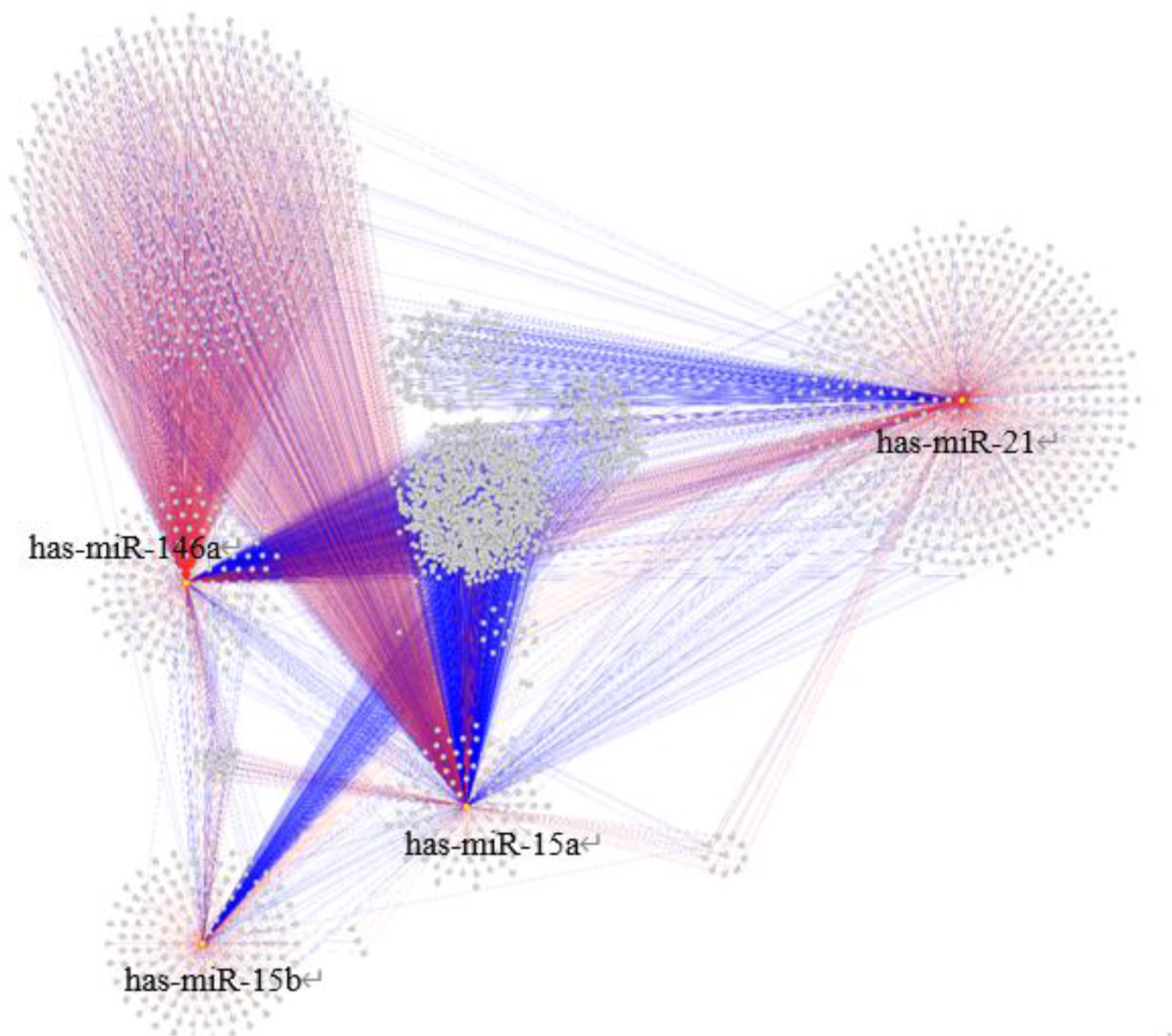
Human GMI (gene-miR interaction) network generated by the CyTargetLinker algorithm human using the miRTarBase (blue) and TargetScan (red) databases. The marked nodes are depicted in yellow. The network is displayed using the yFiles organic layout.

### The concentration-dependent effects of CATs on iPSC-derived cardiomycytess

To investigate the effects of physiological concentrations of CATs (CAT) in TTS patients on the myocardium, we treated human iPSC-derived cardiomycytes with CATs for 24 h. To determine changes in cardiomyocytes upon CAT treatment, we analyzed the expression level of nuclear receptor subfamily 4 group A member 1. This hormone receptor is well expressed in the heart and upregulated after β-adrenergic stimulation.^12^ Additionally to the CATs, cardiomyocytes were treated with a supraphysiological concentration of isoprenaline (Iso). As reported by Borchert et al. the treatment with Iso lead to a significant increase of the cardiac stress marker, whereas physiological CAT concentrations showed no alterations in cardiomyocyte stress levels (Fig. 1a). Multiples of the physiological CAT concentration (10^3^ and 10^6^ of the physiological concentration) lead to cardiac stress indicated by increased expression of NR4A1 (Fig. 1b), 10^9^-fold of the physiological concentration were lethal for the cardiomyocytes.

### Quantification of cytokines and chemokines in patient serum

A previous study demonstrated that treatment with inflammatory cytokines (TNFα, Interleukin-6) leads to compromised endothelial barrier function and reduced or altered junctional protein expression established in vitro model of the BBB, cEND cells ^43^ and in in vivo studies.^44^ Based on these findings, we hypothesized that inflammation is central to the pathophysiology of TTS and explored the time course and persistence of detectable potentially involved pro-inflammatory cytokines. We hypothesized that inflammation is central to the pathophysiology and natural history of Takotsubo syndrome.

We compared the concentration of different cytokines and chemokines in the sera of premenopausal healthy patients, postmenopausal healthy patients, TTS acute, TTS subacute, TTS chronic and ACS (Table 2). Although no statistical significance was found, the concentration of TNFα, IL-2, IL-4 and IL-6 show a tendency to be elevated in the TTS acute group compared to all other groups. In comparison, in the TTS subacute group the concentration of MIP3A, RANTES, EGF, and IL-17 show a tendency to be elevated compared to all other groups. INFγ and IL-10 show higher levels in the postmenopausal healthy group compared to the other groups.

**Table 2.** Measured concentration of different cytokines and chemokines in patients sera in pg/ml ± SD.

|  | premenopausal healthy n = 3 | postmenopausal healthy n= 3 | TTS acute n = 4 | TTS subacute n = 8 | TTS chronic n = 3 | ACS n = 4 |
| --- | --- | --- | --- | --- | --- | --- |
| INFγ | 2,41 ± 1,48 | 18,7 ± 7,18 | 65,6 ± 71,99 | 26,79 ± 32,46 | 19,81 ± 30,50 | 18,46 ± 32,03 |
| IL-2 | 0,32 ± 0,33 | 1,49 ± 1,23 | 4,87 ± 8,012 | 2,2 ± 3,263 | 1,53 ± 2,243 | 0,37 ± 0,3641 |
| IL-4 | 2,34 ± 3,38 | 1,59 ± 0,88 | 50,77 ± 8,68 | 2,75 ± 4,29 | 0,80 ± 1,10 | 0,70 ± 0,92 |
| IL-6 | 0,34 ± 0,34 | 6,64 ± 15,41 | 38,01 ± 45,06 | 8,98 ± 15,13 | 1,45 ± 1,45 | 0,97 ± 0,77 |
| IL-10 | 12,13 ± 19,54 | 30,21 ± 27,33 | 24,48 ± 17,52 | 10,55 ± 10,74 | 3,88 ± 6,68 | 3,41 ± 5,79 |
| TNFα | 15 ± 5,67 | 18,53 ± 18,51 | 41,04 ± 47,72 | 25,8 ± 16,62 | 17,02 ± 5,262 | 15,6 ± 12,43 |
|  |  | postmenopausal healthy n= 3 | TTS acute n = 4 | TTS subacute n = 8 | TTS chronic n = 3 | ACS n = 4 |
| EGF |  | 57,05 | 22,19 | 85,14 | 26,14 | 47,85 |
| IL-17 |  | 3,40 ± 1,70 | 1,91 ± 0,84 | 25,67 ± 52,31 | 3,13 | 3,52 |
| MIP3A |  | 147,3 ± 104,4 | 111,3 ± 68,08 | 249,6 ± 168,5 | 105,3 | 72,69 |
|  |  | postmenopausal healthy n= 3 | TTS acute n = 4 | TTS subacute n = 8 |  |  |
| RANTES |  | 161542 ± 190762 | 69217 ± 81541 | 264518 ± 381721 |  |  |

### Quantification of testosterone and estradiol in serum of patients with various heart diseases and correlation between testosterone-to-estradiol ratio

The sex hormones testosterone and estradiol were measured in serum of healthy postmenopausal patients and patients with TTS (acute, subacute and chronic). The different hormone levels were compared to healthy postmenopausal patients. There was no significant difference between the groups detectable (Table 3).

**Table 3:** Measured concentrations of testosterone and 17ý-estradiol in pg/ml in the serum of patients with various heart disease. Data are the means (± SD) of n independent measures.

|  | postmenopausal healthy n= 3 | TTS acute n = 4 | TTS subacute n = 8 | TTS chronic n = 3 |
| --- | --- | --- | --- | --- |
| Testosterone | 202.5 ± 120.4 | 160 | 287.5 ± 257.5 | 145 ± 63.6 |
| Estradiol | 35.25 ± 54.04 | 5 | 43.13 ± 66.69 | 9.5 ± 6.364 |

The following ratio between testosterone and estradiol (T:E2) as measured in the serum of patients with various heart diseases was calculated: mean T:E postmeno [5.77+0,2]; premeno diestrous [0.5± 0.1]; postmeno estrous [8.1±3.3]; TTS acute [32±0.4]; TTS subacute [6.7±0.1]; TTS chronic [16.11±0.3]. As reference values for healthy premenopausal females the E2-values estrous / di- estrous (400 pg/ml /50 pg/ml/) were used, respectively (University of Rochester, medicine labs).

The calculated T:E2 ratio was the highest in acute TTS serum (32-fold), followed by chronic TTS (26- fold) and subacute TTS (8-fold). Lowest T:E2 ratios were measured in the healthy groups.

### Protein expression of glucocorticoid receptor α in exosomes of serum from patients with various heart diseases

Glucocorticoids are primary stress hormones necessary for life that regulate numerous physiologic processes in an effort to maintain homeostasis.^45^ Glucocorticoids are part of the feedback mechanism in the immune system, which reduces certain aspects of immune function, such as inflammation. The glucocorticoid receptor (GR) is a member of the nuclear hormone receptor superfamily that is activated as a stress-responsive transcription factor upon binding glucocorticoids.^45^ We set out to determine the levels of GRα in patient blood.

For this, after extraction of the exosomes of serum from patients with various heart diseases, Western blot analysis of glucocorticoid receptor α was performed. Changes in protein levels were compared to the healthy postmenopausal group showing a clear trend towards elevation in all TTS group, remarkably the TTS acute and TT chronic groups. Remarkably, even though not significant, values determined from female TTS patients approximate the much higher serum GR levels from male (1,59 of healthy female premenopausal) than as determined from healthy females (postmenopausal healthy set =1; premenopausal healthy = 0.7 ± 0.55).

### Bioinformatic Analysis

Next, we set out to relate the identified inflammatory cytokines present in TTS patient blood to published miRNA in TTS and aging and age-related disease and inflammaging and the CATs described to play a role in the etiology of TTS.^5-8, 22, 23, 46, 47^

A potential miRNA-cytokine-steroid hormone-receptor regulatory network contributing to TTS was developed: The Cytoscape algorithm was able to reconstitute a detailed GMI network comprising 2989 nodes and 6000 edges associated with the corresponding miRs by utilizing the miRTarBase and TargetScan databases (Figure5).^39^ Next, the GGI network was built using the GeneMANIA algorithm for the corresponding blood markers and proinflammatory cytokines comprising 211 nodes and 626 edges (Figure 6).^41^ Finally, the merged network was compiled from the GMI and GGI networks containing 3152 nodes and 5735 edges to connect the blood markers, pro-inflammatory cytokines, and glucocorticoid receptor gene (NR3C1) to miRs (Figure 7 [A]).^40^ This network was processed by subtracting the analyzed nodes to identify only direct gene-miR interactions (Figure 7 [B]). As a result, IL-6, CCL5, CCL20, and INFγ were predicted to be interacted with analyzed miRs directly, while the rest of the genes were involved in indirect interaction with the investigated miRs.

**Figure 6.**
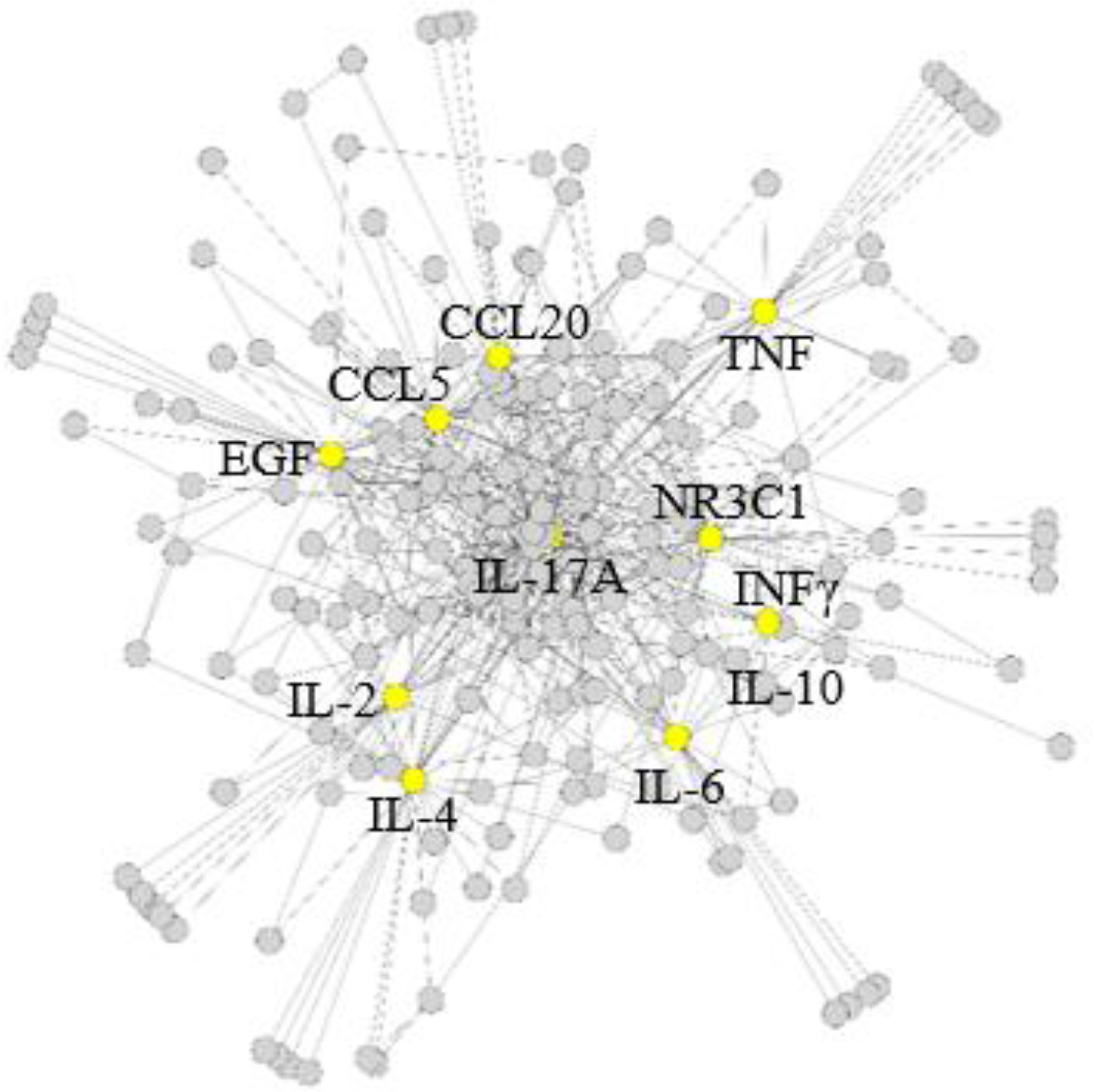
Human GGI (gene-gene interaction) network generated by the GeneMANIA plugin for the corresponding blood markers and proinflammatory cytokines (CCL20, CCL5, EGF, IL2, IL4, IL6, IL10, IL17A, INFγ, NR3C1, and TNF). The marked nodes are depicted in yellow. The network is displayed using the yFiles organic layout.

**Figure 7.**
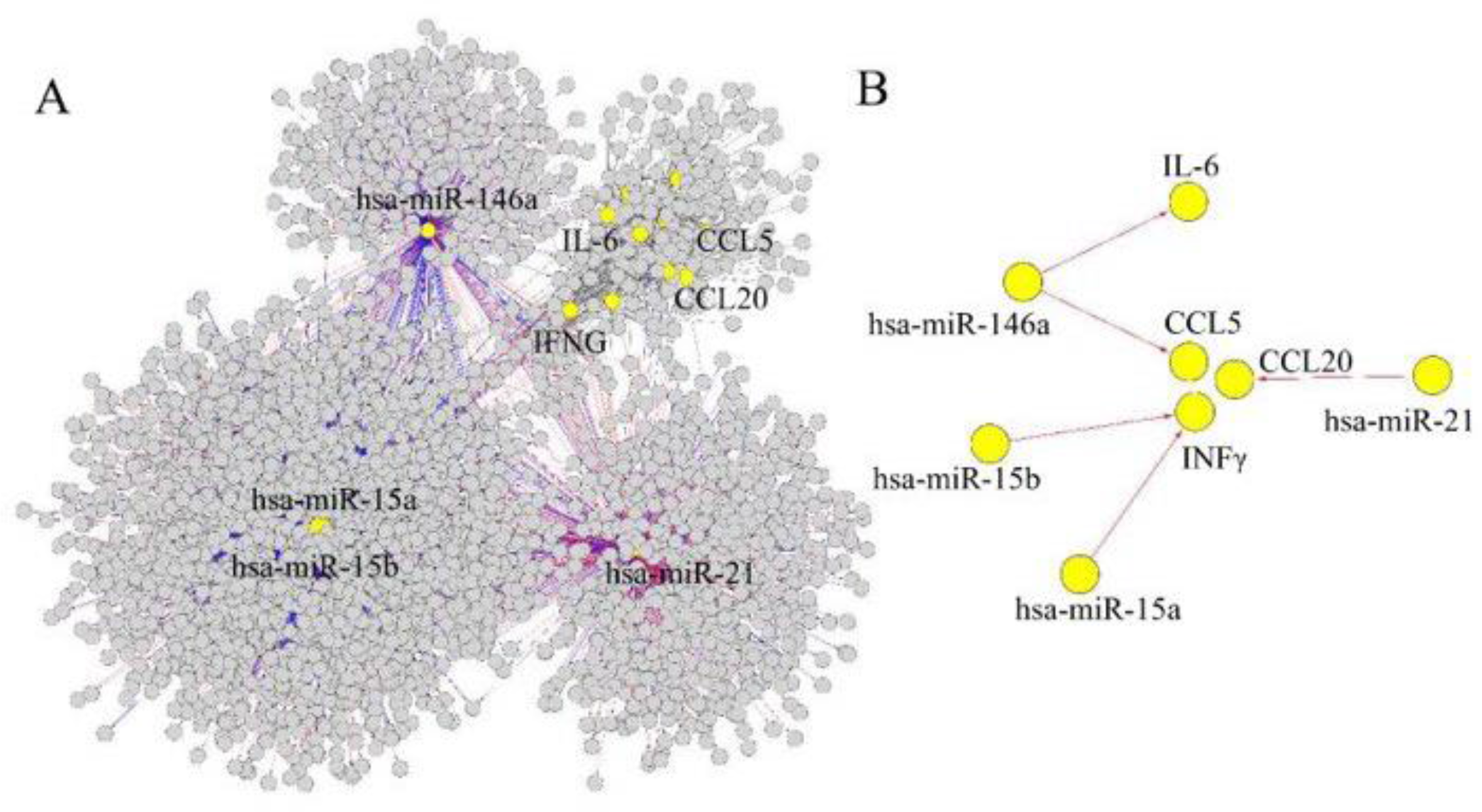
The merged (A) and subtracted (B) networks produced by merging GMI and GGI networks or creating a new network (8 nodes and 5 edges) from selected nodes and all of their edges. The marked nodes are depicted in yellow. The merged network is displayed using the prefuse force- directed layout.

## 4. Discussion

In the clinical practice, a few blood biomarkers are being used to diagnose and monitor TTS relative to the ACS which is characterised by almost the same clinical presentations and ST elevations. These biomarkers serve to predict response to treatment as well as long-term prognosis; blood troponin T, BNP or creatine kinase-MB (CK-MB) are the most frequently surveyed markers.^48, 49^ These markers together with clinicopathological findings ^50, 51^ do convey only insufficient diagnostic accuracy.

### TTS patient serum catecholamine levels

Takotsubo patients present with supraphysiologic plasma CAT levels (epinephrine, norepinephrine, dopamine), which is generally believed to be a cardiotoxic trigger^10, 11^ causal for the abnormal LV contraction pattern in TTS.^10, 11, 52^ Available information remains however controversial^9, 53^ and heavily relies on disease modeling in a dish^30^ or using the isoprenaline rodent model of TTS^38^ creating a supraphysiological CAT exposure for the cell systems or animals investigated while CAT effects on the cardiovascular system in narrower physiological ranges remain elusive. Supporting the CAT hypothesis, some cross-field references can be found in the literature, e.g. pheochromocytoma-related conditions is a CAT-mediated myocarditis and focal/diffuse myocardial fibrosis condition.^54^ Contrast to this hypothesis, our in vitro assessments did not point to structural endothelial cell (from BBB, myocard) or cardiomyocyte damage when exposed to CAT in concentrations as determined from TTS patient blood.

Indirect CAT effects on the cardiovascular system in the pathogenesis of TTS should therefore be considered: meanwhile it has been acknowledged that immune and central nervous systems mutually influence on one another in a bidirectional way, eg in the physiological stress response.^55^ Specifically, the contribution of the locus coeruleus, the principal site for brain synthesis of norepinephrine in the genesis of defeat-stress induced inflammatory signaling should be mentioned.^56^

To conclude, CAT serum levels a reliable TTS biomarker, further research is needed to examine the link between CATs and TTS disease ontogeny. In the following we thus concentrated on an elucidation of disparate novel blood-borne biomarkers of stress and sex differences and inflammation in this stress-related disease.

### TTS patient GR serum levels

We did observe a consistent trend towards elevated GR levels in TTS patient serum. Measured GR serum levels in TTS patient blood clearly exceeded the average postmenopausal healthy female levels 1.6-fold and came close to the values determined from male blood donors.

Generally, stress responses are inhibited by negative feedback mechanisms, whereby glucocorticoids act to diminish drive (brainstem) and promote transsynaptic inhibition by limbic structures (e.g. hippocampus) including feedback downregulation of GR.^57^ In contrast, elevated GR ^58, 59^ expression has been described as a suitable surrogate marker and to positively correlate with glucocorticoid resistance in chronic stress.^60^ Chronic stress-induced activation of the hypothalamus-pituitary-adrenal axis takes many forms (chronic basal hypersecretion, sensitized stress responses, and even adrenal exhaustion), with manifestation dependent upon factors such as stressor chronicity, intensity, frequency, and modality and involves GR insensitivity and upregulation.^57^ Neural mechanisms driving acute and chronic stress responses can be distinct by the serum GR expression, taking into account age, gender and reproductive cycle biases.^61^

Activation of the HPA-axis and sympathetic nervous system overdrive, which are established mechanisms critically involved in the TTS ontogeny are widely accepted to link specifically chronic stress with elevated levels of peripheral pro-inflammatory markers in blood.^62^ Yet, empirical evidence showing that peripheral *levels* of glucocorticoids and/or CATs mediate this effect is equivocal. Recent attention has turned to the possibility that cellular *sensitivity* to these ligands may enhance the effects of inflammatory mediators released under chronic stress.

The glucocorticoid resistance in conjunction with β2-adrenergic receptor signaling pathways in neuronal cells was presented to promote peripheral pro-inflammatory conditions that were postulated to be associated with chronic psychological stress.^63^

However, given the limited size of our study and patient cohort, further research is warranted to examine the link between serum stress, GRα levels and TTS disease progression in female postmenopausal patients.

### Inflammation: cytokines and chemokines in patient serum

The existence of an immune-endocrine relation has been described, pointing out the immuno- modulatory effects of the CATs epinephrine, norepinephrine and dopamine on a wide variety of immune functions.^64^ The authors described that the immunomodulatory effects are mediated by specific adrenergic and dopaminergic receptors detectable on the surface of their immunological target cells and that CATs released within the CNS may chiefly influence the activity of the immune system in the periphery.^64^

We explored expression levels and time course of expression of various pro- and antiinflammatory cytokines while comparing their levels to healthy pre- and postmenopausal subjects plus ACS patients.

Patients with acute TTS had higher serum concentrations of IL-6, CXCL1 (growth-regulated protein) chemokine, TNFα, INFγ, IL-2 and IL-4 compared with control subjects and acute ACS patients, respectively, while EGF-levels were decreased. This observation was in agreement with prior observations published by Scally et al. ^65^ and Santoro,^66^ which also compared inflammatory patterns in TTS and ACS.

Based on the data presented, monitoring the elevation of the cytokines IL-6, TNFα, and INFγ serum levels might be suitable to distinguish TTS acute from ACS, again highlighting the importance of inflammation in the etiology of TTS.

Novel acute phase markers identified include the elevation of both, IL-2 and IL-4. The exploitability of IL-2 (increase) and IL-10 (decrease) as both disease phase markers and distinction markers ACS:TTS needs further exploitation in bigger patient cohorts and warrants future potential as relapse indicators.

The evaluation of IL-2 could represent a novel, TTS acute phase marker, facilitating the discrimination from ACS. Specifically, as IL-2 has been characterized as an anti-inflammatory cytokine, the determination of its levels from patient serum might be a tool to potentially predict propensity to relapse.^67^ However, taking into account the limited number of samples available, a follow-up study including higher sample numbers would be mandatory before final conclusion could be drawn about the suitability of IL-2 as TTS acute phase marker or to differentiate TTS from ACS, or its potential suitability as a relapse marker.

The subacute cytokine profile is most distinctively characterized by elevations in MIP3A, a so far not yet addressed cytokine in frame of TTS, in EGF, TNFα, IL-17 and, interestingly pronounced decreases in IL-10.

Amongst those, IL-17 might represent a future therapeutic target in TTS: Interleukin-17 (IL-17) induces the production of granulocyte colony-stimulating factor (G-CSF) and chemokines such as CXCL1 and CXCL2 and is a cytokine that acts as an inflammation mediator.^68^

Taken together, while the elevation of IL-6, IL-2, IL-4, TNFα, INFγ and CXCL1 can be regarded as TTS acute phase serum markers, a sharp increase in MIP3A and a pronounced decrease in IL-10 was detectable in the TTS subacute, phase which culminated in the chronic phase but also in ACS. In contrast, IL-10 levels were the highest in the postmenopausal healthy group.

### Sex differences

The epidemiological link between female sex and the incidence of TTS cannot be fully explained by stress induced CAT or corticosteroid levels, only. Additional factors, such as female sex or hormone receptor regulation and dysregulated immune response could be an important factor for this specific form of heart failure in postmenopausal patients. Consistent with prior studies, we found strong associations between reduced female sex steroid levels and further extended these studies by calculating an T:E2 ratio which was elevated pronouncedly in TTS patients. Several points could be taken into account:

Women mount stronger immune responses not only against foreign but also against self- antigens, and the prevalence of most autoimmune diseases (AD) is greatly increased in women compared to men. An important role underlying the difference in activity of immune cells in men and women is attributed to sex hormones.^4,5^

Sexually dimorphic actions of glucocorticoids have been shown to provide a link to inflammatory diseases with gender differences in prevalence.^59^

Our data strongly resonate with findings of Walsh et al. ^63^ who report the consistent observation of elevated levels of peripheral pro-inflammatory markers accompanying chronic stress in humans, which associates with not only heightened GR expression, an assumpted result of decreased cellular GR sensitivity, and alterations in ß-AR signaling, which was interpreted to be based on myeloid progenitor cell related stress-signaling pathways.^63^ This situation could also be associated with TTS pathophysiology. Lastly, the exact role that the ratio T:E2 plays in the development of TTS remains unclear and warrants further investigation.

### Bioinformatic extrapolation: TTS – an inflammaging disorder?

The cardiovascular system, and like many other vital organs, the heart is susceptible to the effects of aging. Inflammation is a hallmark of aging, and other coexisting comorbidities linked to age-related decline, such as heart failure, cardiovascular disease, neurodegeneration, vascular dementia and Alzheimer’s disease but also age-related hearing loss, age-related macular degeneration and type 2 diabetes.^24, 69–71^ Inflammation is also associated with TTS. Recently, a novel *portemanteau* of inflammation and aging has been coined, “*inflammaging*”, which is defined as chronic, low-grade inflammation accumulated and worsening over age, presumably contributing to the pathogenesis of various age-related pathologies.^70, 72, 73^ Inflammation has been described to stem from cellular senescence of the immune system, so-called immunosenescence.^74^ Although inflammatory failure,^75^ we set out to lay attention on the role of inflammaging in TTS. In humans and experimental animal models, several reports ^65, 66^ documented a significant association between 4 systemic markers of inflammation, i.e. IL-6, IL-10, TNFα, INFγ.^30, 38, 65, 66^

Our computational analyses predicted IL-6, CCL5, CCL20, and INFγ to interact with the studied miRs directly, while the remaining genes were shown to be involved in indirectly with these miRs. In particular, possible INFγ-signaling pathways were found to be relevant in the ontogeny of TTS (miR- 15a and miR-15b). On the other hand, CCL5, CCL20, and IL-6 were predicted to play a pivotal role in inflammation (miR-21 and miR-146a). Similarly, the IL-6 contribution to the TTS lipidome was previously determined by experimental and computational analyses connecting this cytokine to the phosphatidylinositol molecule.^76^

Currently, the role of inflammaging in the ontogeny of TTS is only speculative, and further large-scale studies are needed to verify.

To conclude, the contributions of various molecular mechanisms, notably inflammaging, oxidative stress, CUMS, gender, and genetic factors shall be evaluated as possible common culprits that interrelate the pathophysiology in the heart and cardiovascular system as part of TTS. Future research using new technologies like big-data analysis of blood samples from real-life patient cohorts will probably gain better insight into underlying mechanisms of different disease phenotypes. Identification of specific molecular pathways and associated biomarkers will then allow the development of new targets for precision medicine.

## Data Availability

The original contributions presented in this study are included in the article/supplementary material, further inquiries can be directed to the corresponding author.

## Acknowledgment

The assistance provided by Elisabeth Wilken and Mariola Dragan was greatly appreciated. We wish to extend special thanks to Hiroshima Asa Citizens hospital team for technical support. The authors wish to show their appreciation to Dr. C. Maack, Comprehensive heart failure center Würzburg, Germany for inspiring scientific discussions. The research grant DFG FO 315/5-1 to CF is acknowledged.

## Authors’ Contributions

MS and HH performed experiments, conducted data and statistical analyses, and prepared the manuscript. SS performed computational analyses. MN, CF and SS, CB, HH prepared the manuscript. CF, CB and TT conceptualized the project and supervised the experiments. All authors contributed to manuscript writing and proofreading.

## Abbreviations

ACS: acute coronary syndrome; AD: autoimmune disease; BBB: blood-brain barrier; b-FGF: basic fibroblast growth factor; BNP: brain natriuretic peptide; CAT: cathecholamine; CCL: CC-chemokine ligand; cEND: mouse brain capillary endothelial cell line; CK-MB: creatine kinase MB; CM: cardiomyocyte; CNS: Central Nervous System; CUMS: chronic unpredictable mild stress; DMEM: Dulbecco’s modified Eagle’s medium; ECG: electrocardiogram; EGF: epidermal growth factor; FCS: fetal calf serum; G-CSF: granulocyte colony-stimulating factor; GGI: gene-gene interaction; GMI: gene-miR interaction; GR: glucocorticoid receptor; GSK-3: Glycogen synthase kinase 3; GUSB: β-glucuronidase; hCMEC/D3: human brain vascular endothelial cell line; HPA: human platelet antigen; IL: interleukin ; INFγ: Interferon gamma; iPSC: induced pluripotent stem cells; iPSC-CMs: induced pluripotent stem cells differentiated into cardiomyocytes; Iso: isoprenaline; LV: left ventricle; MIP3A: Macrophage Inflammatory Protein 3 alpha; miR/miRNA: microRNA; MyEND: myocardial endothelial cell line; NEA: non-essential amino acids; NR3C1: nuclear receptor subfamily 3 group C member 1; NR4A1: nuclear receptor subfamily 4 group A member 1; P/S: penicillin/streptomycin; PET: positronenemissionstomography; RT-qPCR: real-time quantitative PCR; R3-IGF-1: recombinant analog of insulin-like growth factor; RANTES: CC-Chemokin-Ligand- 5; SD: standard deviation; STEMI: ST-segment elevation myocardial infarction; TNFα: tumor necrosis factor-α; TTC: takotsubo cardiomyopathy; TTS: Takotsubo syndrome; VEGF: Vascular Endothelial Growth Factor; ZO-1: Zonula Occludens 1; β2-AR: β2-adrenergic receptor

